# Clonal hematopoiesis is associated with risk of severe Covid-19

**DOI:** 10.1101/2020.11.25.20233163

**Authors:** Kelly L. Bolton, Youngil Koh, Michael B. Foote, Hogune Im, Justin Jee, Choong Hyun Sun, Anton Safonov, Ryan Ptashkin, Joon Ho Moon, Ji Yeon Lee, Jongtak Jung, Chang Kyung Kang, Kyoung-Ho Song, Pyeong Gyun Choe, Wan Beom Park, Hong Bin Kim, Myoung-don Oh, Han Song, Sugyeong Kim, Minal Patel, Andriy Derkach, Erika Gedvilaite, Kaitlyn A. Tkachuk, Lior Z. Braunstein, Teng Gao, Elli Papaemmanuil, N. Esther Babady, Melissa S. Pessin, Mini Kamboj, Luis A. Diaz, Marc Ladanyi, Michael J. Rauh, Pradeep Natarajan, Mitchell J. Machiela, Philip Awadalla, Vijai Joseph, Kenneth Offit, Larry Norton, Michael F Berger, Ross L Levine, Eu Suk Kim, Nam Joong Kim, Ahmet Zehir

## Abstract

Acquired somatic mutations in hematopoietic stem and progenitor cells (clonal hematopoiesis or CH) are associated with advanced age, increased risk of cardiovascular and malignant diseases, and decreased overall survival.^1–4^ These adverse sequelae may be mediated by altered inflammatory profiles observed in patients with CH.^2,5,6^ A pro-inflammatory immunologic profile is also associated with worse outcomes of certain infections, including SARS-CoV-2 and its associated disease Covid-19.^7,8^ Whether CH predisposes to severe Covid-19 or other infections is unknown. Among 515 individuals with Covid-19 from Memorial Sloan Kettering (MSK) and the Korean Clonal Hematopoiesis (KoCH) consortia, we found that CH was associated with severe Covid-19 outcomes (OR=1.9, 95%=1.2-2.9, p=0.01). We further explored the relationship between CH and risk of other infections in 14,211 solid tumor patients at MSK. CH was significantly associated with risk of *Clostridium Difficile* (HR=2.0, 95% CI: 1.2-3.3, p=6×10^−3^) and *Streptococcus/Enterococcus* infections (HR=1.5, 95% CI=1.1-2.1, p=5×10^−3^). These findings suggest a relationship between CH and risk of severe infections that warrants further investigation.

## MAIN

Acquired mutations that lead to clonal expansion are common in the normal aging hematopoietic system (clonal hematopoiesis, or CH), yet are known to alter stem/progenitor and lymphoid function and response to environmental stressors, including systemic infections^5,6,9,10^. The mutational events that drive CH overlap with known drivers of hematologic malignancies. However, the majority of mutations in CH appear to occur outside of canonical cancer driver genes^11,12^. The impact of individual mutational events on hematopoietic stem and progenitor cells differs by the nature of the genomic aberration. For example, chromosomal aneuploidies result in a predisposition for lymphoid fate specification and transformation^13,14^ while point mutations in *DNMT3A* result in increased myeloid differentiation^6,15^. Heterogeneity also exists across CH phenotypes by driver gene in regards to its impact on inflammatory signaling^6^. For example, mutations in *TET2* result in heightened secretion of several cytokines including IL-1β/IL-6 signaling that may partially explain the increased risk of cardiovascular disease^5,9,16^. Moreover, systemic infections and the resultant inflammatory signals can lead to increased clonal fitness of *TET2* mutant cells and clonal expansion^10,17,18^.

Despite these important insights, the relationship between different CH events, infectious risk and infectious disease severity has not been studied. The severity of Covid-19 is also associated with advanced age, cardiovascular and malignant comorbidities, and elevated circulating IL-6 levels; features which are seen with age-associated CH^19–23^. Given the common inflammatory profile of CH and Covid-19 infection, we investigated the relationship between CH and Covid-19 including the potential for an association of CH with increased Covid-19 disease severity.

Our study included patients from two separate cohorts. The first cohort was composed of patients with solid tumors treated at Memorial Sloan Kettering Cancer Center (MSK) with blood previously sequenced using MSK-IMPACT, a previously validated targeted gene panel capturing all commonly mutated CH-associated genes (Supplementary Table 1)^24^. Of these patients, 1,626 were tested for SARS-CoV-2 (the virus that causes Covid-19) RNA between March 1st 2020 and July 1st 2020; 403 (24.8%) individuals tested positive for SARS-CoV-2 (Methods and Table 1). The second cohort included 112 previously healthy individuals without cancer who were hospitalized for Covid-19 between January and April 2020 at four tertiary hospitals in South Korea (KoCH cohort). The KoCH cohort was sequenced using a custom targeted NGS panel from Agilent (89 genes) which was designed to include commonly occurring CH genes (Supplementary Table 2).

**Table 1.** Characteristics of study participants.

|  | MSK |  |  |  | KoCH |  |
| --- | --- | --- | --- | --- | --- | --- |
|  | Severe Covid<br>(N=94) | Non-severe Covid<br>(N=319) | Negative<br>(N=1223) | Untested<br>(N=7681) | Severe Covid<br>(N=68) | Non-Severe Covid<br>(N=44) |
| **Age(y)** |  |  |  |  |  |  |
| 0-50 | 28 (17.1%) | 70 (25.9%) | 302 (25.2%) | 1703 (22.3%) | 6 (8.8%) | 16 (36.4%) |
| 50-60 | 32 (19.5%) | 82 (30.4%) | 297 (24.8%) | 1731 (22.6%) | 10 (14.7%) | 6 (13.6%) |
| 60-70 | 51 (31.1%) | 74 (27.4%) | 348 (29.1%) | 2348 (30.7%) | 23 (33.8%) | 8 (18.2%) |
| 70-80 | 38 (23.2%) | 39 (14.4%) | 206 (17.2%) | 1518 (19.8%) | 15 (22.1%) | 9 (20.5%) |
| 80+ | 15 (9.1%) | 5 (1.9%) | 44 (3.7%) | 348 (4.6%) | 14 (20.6%) | 5 (11.4%) |
| **Gender** |  |  |  |  |  |  |
| Female | 89 (54.3%) | 144 (53.3%) | 662 (55.3%) | 4546 (59.4%) | 28 (41.2%) | 23 (52.3%) |
| Male | 75 (45.7%) | 126 (46.7%) | 535 (44.7%) | 3102 (40.6%) | 40 (58.8%) | 21 (47.7%) |
| **Smoking** |  |  |  |  |  |  |
| never | 71 (43.3%) | 130 (48.1%) | 556 (46.4%) | 3670 (48.0%) | 34 (50.0%) | 30 (68.2%) |
| current_former | 88 (53.7%) | 134 (49.6%) | 622 (52.0%) | 3894 (50.9%) | 10 (14.7%) | 13 (29.5%) |
| missing | 5 (3.0%) | 6 (2.2%) | 19 (1.6%) | 84 (1.1%) | 24 (35.3%) | 1 (2.3%) |
| **Hypertension** |  |  |  |  |  |  |
| No | 74 (45.1%) | 158 (58.5%) | 660 (55.1%) | 4400 (57.5%) | 34 (50.0%) | 26 (59.1%) |
| Yes | 90 (54.9%) | 112 (41.5%) | 537 (44.9%) | 3248 (42.5%) | 34 (50.0%) | 18 (40.9%) |
| **Coronary Artery Disease** |  |  |  |  |  |  |
| No | 140 (85.4%) | 244 (90.4%) | 1078 (90.1%) | 7032 (91.9%) | 63 (92.6%) | 40 (90.9%) |
| Yes | 24 (14.6%) | 26 (9.6%) | 119 (9.9%) | 616 (8.1%) | 5 (7.4%) | 4 (9.1%) |
| **COPD/Asthma** |  |  |  |  |  |  |
| No | 133 (81.1%) | 233 (86.3%) | 1005 (84.0%) | 6505 (85.1%) | 64 (94.1%) | 41 (93.2%) |
| Yes | 31 (18.9%) | 37 (13.7%) | 192 (16.0%) | 1143 (14.9%) | 4 (5.9%) | 3 (6.8%) |
| **Diabetes** |  |  |  |  |  |  |
| No | 120 (73.2%) | 221 (81.9%) | 1019 (85.1%) | 6657 (87.0%) | 48 (70.6%) | 36 (81.8%) |
| Yes | 44 (26.8%) | 49 (18.1%) | 178 (14.9%) | 991 (13.0%) | 20 (29.4%) | 8 (18.2%) |
| **Race** |  |  |  |  |  |  |
| white | 91 (55.5%) | 175 (64.8%) | 890 (74.4%) | 5701 (74.5%) | 0(0.0%) | 0(0.0%) |
| missing | 11 (6.7%) | 18 (6.7%) | 53 (4.4%) | 327 (4.3%) | 0(0.0%) | 0(0.0%) |
| non-white | 62 (37.8%) | 77 (28.5%) | 254 (21.2%) | 1620 (21.2%) | 68 (100%) | 44 (100%) |
| **Cytotoxic Therapy Prior to Blood Draw** |  |  |  |  |  |  |
| No | 122 (74.4%) | 194 (71.9%) | 727 (60.7%) | 5538 (72.4%) |  |  |
| Yes | 42 (25.6%) | 76 (28.1%) | 470 (39.3%) | 2110 (27.6%) |  |  |
| **Cytotoxic Therapy After Blood Draw** |  |  |  |  |  |  |
| No | 46 (28.0%) | 122 (45.2%) | 345 (28.8%) | 4198 (54.9%) |  |  |
| Yes | 118 (72.0%) | 148 (54.8%) | 852 (71.2%) | 3450 (45.1%) |  |  |
| **Primary Tumor Site** |  |  |  |  |  |  |
| Other primary tumor site | 123 (75.0%) | 224 (83.0%) | 909 (75.9%) | 6072 (79.4%) |  |  |
| Thoracic primary tumor site | 41 (25.0%) | 46 (17.0%) | 288 (24.1%) | 1576 (20.6%) |  |  |

For both cohorts, the primary outcome was severe Covid-19 infection, defined as the presence of hypoxia requiring supplemental oxygen (oxygen device >1 L or hypoxia <94%). We used multivariable logistic regression adjusting for covariates including age, smoking, prior Covid-19 related comorbidities, and prior cancer treatment to determine the association between severe Covid-19 and CH in each population. We then performed a fixed-effects meta-analysis to estimate the association in the overall population. The full statistical rationale is further described in the Methods section.

Among Covid-19 positive individuals, 23% (N=94) and 61% (N=68) had severe disease in the MSK and KoCH cohorts, respectively (Table 1). Overall, CH was observed in 35% of Covid-19 positive cases at MSK and 21% in KoCH. Of note, when restricting the MSK-IMPACT panel to the 89 genes included in the KoCH panel, 20% of Covid-19 positive cases at MSK had CH. In the MSK cohort, CH was observed in 51% and 30% of patients with severe versus non-severe Covid-19, respectively (adjusted OR: 1.85, 95% CI 1.10-3.12, Figure 1). In the KoCH cohort, CH was observed in 25% and 15.9% of patients with severe versus non-severe Covid-19, respectively (adjusted OR 1.85, 95% CI 0.53-6.43, Figure 1). In a fixed effects meta-analysis of odds-ratio estimates from the multivariable logistic regression models employed in each separate cohort analysis, the presence of CH was associated with an increased risk of severe Covid-19 (OR=1.85, 95%=1.15-2.99, p=0.01) (Figure 1).

**Figure 1.**
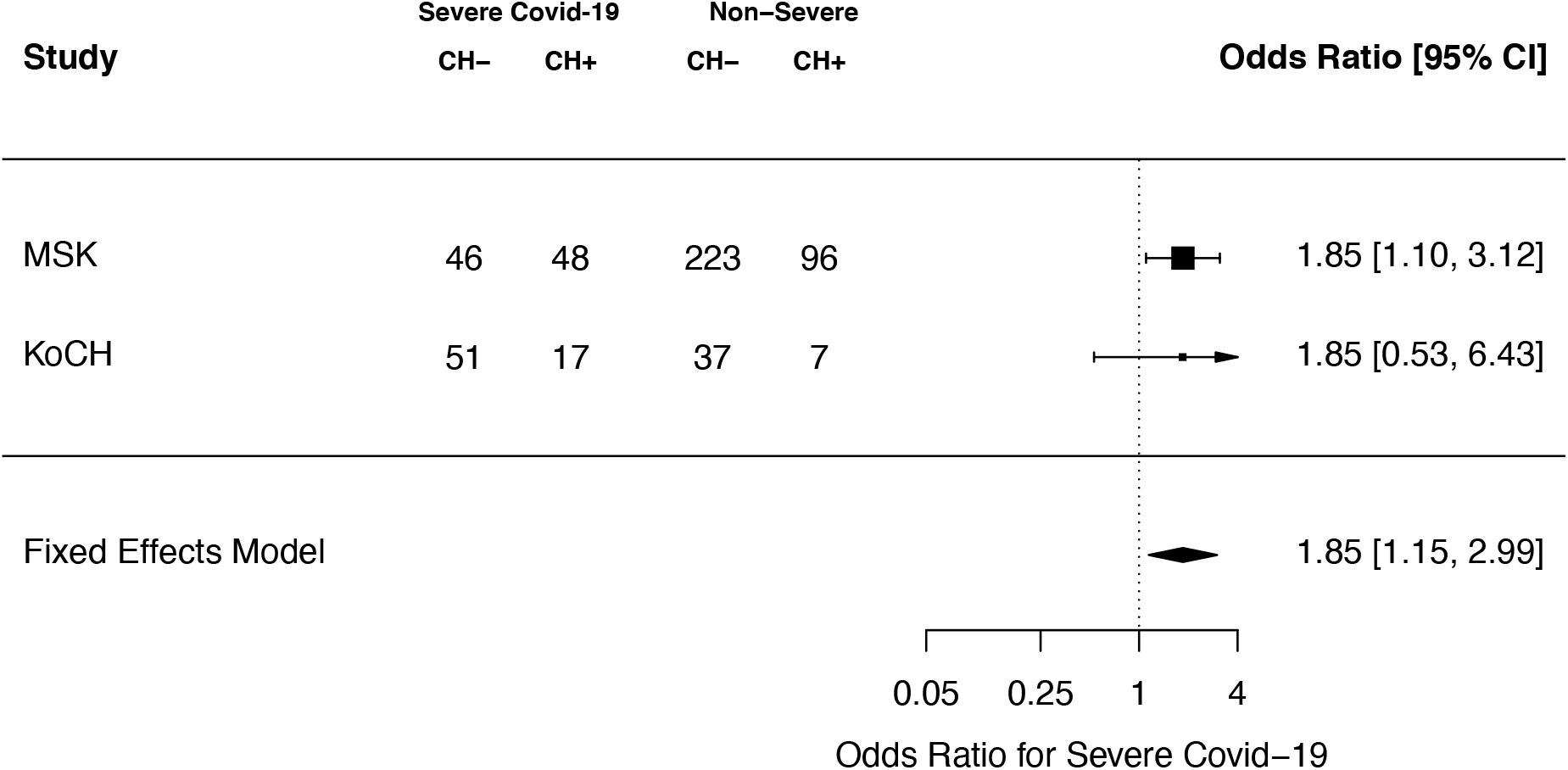
Association between CH and Covid-19 severity. Shown are the results from logistic regression adjusted for age, gender, race, smoking, diabetes, cardiovascular disease, COPD/asthma, cancer primary site (if history of malignancy), exposure to cytotoxic cancer therapy for the MSK and KoCH. Summary statistics for a fixed effects meta-analysis are shown.

Using previously described methods^24^, CH mutations were classified as known or hypothesized cancer putative drivers (PD-CH) or non putative drivers (non-PD CH). The majority of CH mutations were classified as PD-CH (52% in the MSK cohort and 67% in the KoCH dataset). In order to explore the association between particular mutation types and Covid-19 severity, we performed a stratified analysis of Covid-19 severity by PD-CH versus non-PD CH status. A significant association was observed between non-PD CH and severe Covid-19 (OR=2.01, 95% CI=1.15-3.50, p=0.02), as well as between silent (synonymous) CH and severe Covid-19 (OR=2.58, 95% CI 1.01-6.61, p=0.05). There was not a statistically-significant association between PD-CH and severe Covid-19 infection (OR=1.15, 95% CI=0.61-2.02, p=0.77: Extended Data Figure 1). Most non-PD mutations in Covid-19 positive cases occurred in non-recurrently mutated genes (65% at MSK and 76.9% in KoCH, Supplementary Figure 1).

The strength of the association between CH and severe Covid-19 was similar among patients with one CH mutation (OR=1.78, 95% CI=1.0-3.1, p=0.04) and multiple CH mutations (OR=2.0, 95% CI=1.0-3.8, p=0.04). Patients with a maximum CH variant allele frequency (VAF) of >5% showed a significant association with severe Covid-19 (OR=1.9, 95% CI=1.0-3.4, p=0.04). This association trended towards statistical significance in patients with any CH mutation and a maximum VAF<5% (OR=1.75, 95% CI=0.97-3.17, p=0.06: Extended Data Figures 2-4). These data suggest that the presence of CH and resultant alterations in hematopoietic differentiation, and not specific mutant alleles, is predictive of Covid-19 disease severity.

Given the evidence of an association between CH and Covid-19 severity, we sought to explore the relationship between CH and other types of infections. We analyzed billing codes from 14,211 solid tumor patients treated at MSK who underwent blood sequencing by MSK-IMPACT. Using a previously established phenome-wide-association study (Phe-WAS) methodology^25^, we mapped patient ICD-9 and ICD-10 billing codes to categories of infectious disease. Multivariable Cox proportional hazards regression was used to estimate the hazard ratio (HR) for risk of infection among CH positive compared to CH negative individuals. Given the number of model covariates, we limited the analysis to 32 infection subclasses that affected at least 80 individuals (see Methods). Multiple infection types were associated with CH, although many associations were not statistically-significant after multiplicity adjustment (Figure 2A, Supplementary Table 3). CH was significantly (FDR-corrected p-value<0.10) associated with the onset of two infection subclasses: *Clostridium difficile* infection (HR=2.0, 95% CI: 1.2-3.3, p=6×10^−3^) and *Streptococcus/Enterococcus* infection (HR=1.5, 95% CI=1.1-2.1, p=5×10^−3^). When stratified by CH-mutation characteristics, patients with two or more CH-mutations had a stronger association with *Clostridium difficile* infection (OR=3.4, 95% CI=1.8-6.3, p=2×10-4) compared to patients with one CH-mutation (OR=1.4, 95% CI=0.8-2.7, p=0.28). The association between CH and *Clostridium difficile* infection was significant for mutations with a VAF of >5% (OR=2.5, 95% CI=1.4-4.6, p=0.002) but not mutations with a VAF of 2-5% (OR=1.6, 95% CI=0.8-3.1, p=0.17). Similar to Covid-19 severity, the association between CH and *Clostridium difficile* infection was significant for non-PD CH (OR=2.0, 95% CI=1.2-3.3, p=0.01) and silent mutations (OR=2.6, 95% CI=1.2-5.8, p=0.02) but not CH-PD (OR=1.4, 95% CI=0.7-2.8, p=0.39) (Figure 2B).

**Figure 2.**
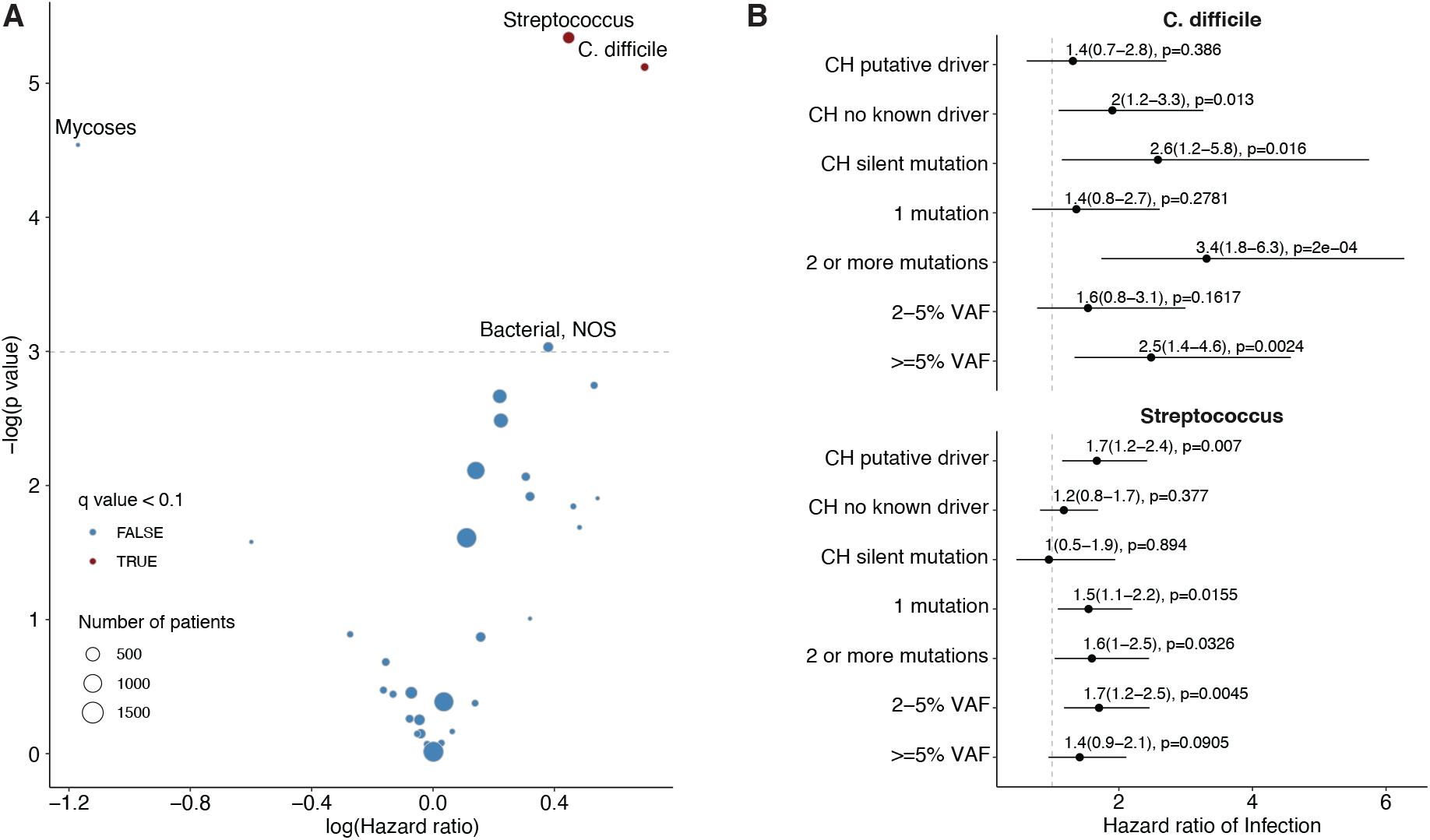
Association between CH and risk of infection in solid tumor patients. A) Volcano plot of the log(Hazard ratio) of infection with CH using multivariable cox proportional hazards regression. B) Association between CH subtype defined by putative driver status and risk of *Clostridium Difficle* and *Streptococcus/Enterococcus* infection using cox proportional hazards regression. All models were adjusted for age, gender, race, smoking, diabetes, cardiovascular disease, COPD/asthma, cancer primary site (if history of malignancy), exposure to cytotoxic cancer therapy.

In summary, we show in cancer and non-cancer patients that CH is associated with increased Covid-19 severity. In a large cancer patient cohort, CH is also associated with other severe infections, namely *Streptococcus/Enterococccus* and *Clostridium difficile* infections. Our exploratory analysis suggests that the relationship between CH and Covid-19 and CH and *Clostridium* difficile infection may be partly driven by non-driver CH. Clonal expansions characterized by non-driver mutational events could be facilitated by multiple mechanisms. Many classes of genetic alterations, such as copy number events (CNVs), structural variants, non-coding, and epigenetic changes, are not detectable using the targeted panels included in this study. As such, the observed events that are highly enriched in CH could be ‘passenger’ mutations that co-occur with a positively selected, undetected ‘driver’ mutation such as recurrent CNVs.^12,13^ Alternatively, driver mutations may have been incompletely classified as “non-driver” events using our methodology. However, cancer driver genes tend to recur in multiple patients, and the majority of witnessed non-driver mutated genes in our cohort were non-recurrent suggesting that clonal expansion, and not the specific event driving clonal expansion, may be associated with Covid-19 disease severity. This will need to be further studied in larger cohorts.

The hematopoietic system is a key regulator of inflammation and immunity. A substantial body of evidence now links somatic alterations in hematopoietic stem and progenitor cells to a variety of health outcomes, with inflammation emerging as a key mediator.^2–5,10–13^ Our data, along with results in the accompanying manuscript by Zekavat et al., demonstrate a similar association between CH and increased infection severity. This association may be due to residual confounding by variables that are unknown and unaccounted for in our models. Alternatively, this could represent a novel pathophysiology that links CH-induced changes in hematopoietic stem, progenitor, and lymphoid cell function with immune regulation and infection response. Future investigation including functional studies will be important to clarify the mechanisms underlying the association between CH and infection risk and to develop potential interventional strategies to attenuate inflammation, clonal expansion, and infectious sequelae in patients with and without cancer.

## METHODS

### Sample ascertainment and clinical data extraction

#### MSK Cohort

The study population included 9,307 patients with non-hematologic cancers at MSKCC who were alive on May 1st 2020 and underwent matched tumor and blood sequencing before this date using the MSK-IMPACT panel on an institutional prospective tumor sequencing protocol (ClinicalTrials.gov number, NCT01775072). Subjects who had a hematologic malignancy diagnosed after MSK-IMPACT testing or who had an active hematologic malignancy at the time of blood draw were excluded. Demographics, smoking history, exposure to oncologic therapy and primary tumor site were extracted from the electronic health record. Accuracy of populated information was manually checked in the EMR by three independent physicians (K.B, M.F, A.S). The presence of co-existing medical comorbidities known to correlate with Covid-19 severity including diabetes, COPD, asthma, hypertension and cardiovascular disease, were ascertained from ICD-9 and ICD-10 billing codes. SARS-CoV-2 status was determined using RT-PCR. We defined severe Covid-19 as the presence of hypoxia requiring supplemental oxygen (supplemental oxygen device >1 L or hypoxia <94%) resulting from Covid-19 infection. There were seven subjects with Covid-19 for whom there was minimal documentation of clinical course following Covid-19 infection and these individuals were excluded. There were three individuals with metastatic cancer and progression of disease at the time of Covid-19 where it was unclear whether documented hypoxia could be attributed to Covid-19 or disease progression. These subjects were also excluded.

#### KoCH Cohort

Laboratory-confirmed patients with Covid-19 between January and April 2020 in four tertiary hospitals in Republic of Korea were approached for consent to this study. Blood was drawn following confirmation of Covid-19 positivity. All of four hospitals have been running national-designated isolation units, which are located in Seoul, Gyeonggi, or Daegu. These provinces had the highest numbers of Covid-19 cases during the period^14^. Clinical and laboratory characteristics were retrospectively reviewed using the electronic medical record systems of each institution. Hypoxia requiring supplemental oxygen was defined as supplemental oxygen device >1 L with O2 <94% resulting from Covid-19 infection. Institutional Review Boards (IRBs) of four respective hospitals approved the study (IRB No. 2003-141-1110, B-2006/616-409), and written informed consents of the patients were obtained per IRB recommendations. Subjects who had an active malignancy at the time of blood draw were excluded.

### Sequencing and Variant Calling

#### MSK Cohort

Subjects had a tumor and blood sample (as a matched normal control) sequenced using MSK-IMPACT, an FDA-authorized hybridization capture-based next-generation sequencing assay encompassing all protein-coding exons from the canonical transcript of 341, 410, or 468 cancer-associated genes (Supplementary Table 4). MSK-IMPACT is validated and approved for clinical use by New York State Department of Health Clinical Laboratory Evaluation Program. The sequencing test utilizes genomic DNA extracted from formalin fixed paraffin embedded (FFPE) tumor tissue as well as matched patient blood samples. DNA is sheared and DNA fragments are captured using custom probes^47^. MSK-IMPACT contains most of the commonly reported CH genes with the exception that earlier versions of the panel did not contain *PPM1D* or *SRSF2*.

Pooled libraries were sequenced on an Illumina HiSeq 2500 with 2×100bp paired-end reads. Sequencing reads were aligned to the human genome (hg19) using BWA (0.7.5a). Reads were re-aligned around indels using ABRA (0.92), followed by base quality score recalibration with Genome Analysis Toolkit (GATK) (3.3-0). Median coverage in the blood samples was 497x, and median coverage in the tumors was 790x. Variant calling for each blood sample was performed unmatched, using a pooled control sample of DNA from 10 unrelated individuals as a comparator. Single nucleotide variants (SNVs) were called using Mutect and VarDict. Insertions and deletions were called using Somatic Indel Detector (SID) and VarDict. Variants that were called by two callers were retained. Dinucleotide substitution variants (DNVs) were detected by VarDict and retained if any base overlapped a SNV called by Mutect. All called mutations were genotyped in the patient-matched tumor sample. Mutations were annotated with VEP(version 86) and OncoKb. We applied a series of post-processing filters to further remove false positive variants caused by sequencing artifacts and putative germline polymorphisms as previously described^24^.

#### KoCH Cohort

Blood-derived DNA was sequenced using a custom panel of 89 genes frequently mutated in CH. All NGS libraries were prepared using the Agilent SureSelect XT HS and XT Low input enzymatic fragmentation kit. Pooled Libraries were sequenced on an Illumina NovaSeq6000 with 2×150bp paired-end reads. Sequencing reads were trimmed with SeqPrep (v0.3) and Sickle (v1.33) and aligned to the human genome (hg19) using BWA-MEM (v0.7.10). PICARD(v1.94) was used for duplicate marking followed by indel realignment and base quality score recalibration with GATK light(v2.3.9). The mean depth of coverage of samples was higher than 800x. Variant calling was performed using SNver(v0.4.1), LoFreq(v0.6.1), GATK UnifiedGenotyper(v2.3.9) for SNVs. For Insertions and deletions in-house InDel caller was used^26^. The union of all called results were filtered meeting the internal criteria (total reads >=10, Alt reads >=10, positive Alt reads >=5, negative Alt reads >=5, MQV >=30, BQV >=30) and VAF falling between 2% and 30%. Common germline variants were filtered based on genomAD, 1k Genome v3, ESP6500 and ExAC data. Lastly, technical artifact calls with maf >2% were filtered based on an internal panel of 1000 individuals who were CH negative.

### Variant Annotation

Variants from the MSK and KoCH cohort were uniformly annotated according to evidence for functional relevance in cancer (putative driver or CH-PD). We annotated variants as oncogenic if they fulfilled any of the following criteria: 1) truncating variants in *NF1, DNMT3A, TET2, IKZF1, RAD21, WT1, KMT2D, SH2B3, TP53, CEBPA, ASXL1, RUNX1, BCOR, KDM6A, STAG2, PHF6, KMT2C, PPM1D, ATM, ARID1A, ARID2, ASXL2, CHEK2, CREBBP, ETV6, EZH2, FBXW7, MGA, MPL, RB1, SETD2, SUZ12, ZRSR2* or in *CALR* exon 9; 2) any truncating mutations (nonsense, essential splice site or frameshift indel) in known tumor suppressor genes as per the Cancer Gene Census, OncoKB, or the scientific literature; 3) translation start site mutations in *SH2B3*; 4) *TERT* promoter mutations; 5) *FLT3*-ITDs; 6) in-frame indels in *CALR, CEBPA, CHEK2, ETV6, EZH2*; 7) any variant occurring in the COSMIC “haematopoietic and lymphoid” category greater than or equal to 10 times; 8) any variant reported as somatic at least 20 times in COSMIC; 9) any variant noted as potentially oncogenic in an in-house dataset of 7,000 individuals with myeloid neoplasm greater than or equal to 5 times; 10) any loci (defined by the amino acid location) reported as having at least 5 missense mutations and at least one exact mutational match in TopMed^6^.

### Statistical analysis

#### CH and Covid-19 Severity

We used multivariable logistic regression to evaluate for an association between clonal hematopoiesis and Covid-19 severity adjusting for age (measured as a continuous variable), gender, race, smoking history and co-existing medical comorbidities including diabetes, COPD/asthma and cardiovascular disease all classified as per Table 1. This was done separately for the MSK and KoCH cohorts due to limitations of data sharing. For solid tumor patients at MSK we also adjusted for primary tumor site (thoracic or non-thoracic cancer) and receipt of cytotoxic chemotherapy before and after IMPACT blood draw. We performed a fixed effects meta-analysis of the MSK and KoCH cohorts to jointly estimate the odds ratio for severe Covid-19 among CH positive compared to CH negative individuals.

#### CH and risk of infection in the MSK cohort

We analyzed billing codes from 14,211 solid tumor patients at MSKCC who had their blood sequenced using MSK-IMPACT. We applied the phecode nomenclature developed at Vanderbilt^9^ to map ICD-9 and ICD-10 billing codes to infectious disease subtypes. Subjects who were billed using a ICD9/10 code within the phecode for the first time following their sequencing blood draw with evidence of CH were considered to have an incident infection. Those who were billed for an ICD9/10 code within the phecode prior to blood draw were removed from the analysis of that phecode. In order to evaluate the accuracy of the billing code data, the presence of a documented *Clostridium Difficile* or *Streptococcus* infection in an EMR physician note was manually checked for patients respectively identified by billing codes (N=525 patients) by three independent physicians using shared criteria for infection onset. Billing codes were highly accurate in identifying the presence of the respective infectious disease (concordance >95%).

We used Cox proportional hazards regression to estimate the hazard ratio for risk of infection among those with CH compared to CH negative individuals. The date of blood draw (used for MSK-IMPACT sequencing) served as the onset date for this time-to-event analysis; the end-date was the date of billing code entry for the infectious disease subtype phecode, death or last follow-up, whichever came first. All models were adjusted for age, gender, race, smoking, tumor type, and cumulative exposure to cytotoxic chemotherapy prior to blood draw and after blood draw as previously described^10^. Following the 10:1 rule regarding the number of covariates in a multivariable model in proportion to the number of events^16^, we excluded infection subclasses populated with less than 80 individuals. The analysis utilized multiplicity correction with the Benjamini-Hochberg method to establish adjusted q-values for hazard ratio with a prespecified false-discovery-rate (FDR) <0.10.

All the statistical analyses were performed with the use of the R statistical package (www.r-project.org).

## Data Availability

The minimal clinical and genetic data frame used to generate all analyses presented in this paper are available by request and will be made publicly available upon acceptance to a peer-reviewed journal.

## Author Contributions

K.L.B, M.F, J.J, Y.K, H.I, C.S, E.K, N.K, A.Z conceived and designed the study. K.L.B, M.F., J.J., A.S, J.M, J.L, J.J, C.K, K.S, P.C, W.P, H.K, M.O, H.S, S.K,E.K, N.K performed collection of clinical data. K.L.B, R.P, T.G, A.S, M.B,Y.K,H.I,C.S, A.Z led the generation and analysis of sequencing data. K.L.B, M.F, J.J., A.S., Y.K, H.I, C.S, S.K,H.S,A.Z, performed statistical analyses and/or participated in data interpretation. All authors contributed to the writing of the manuscript and approved it for submission.

## Acknowledgements

This work was supported by the National Institute of Health (K08CA241318 to K.L.B, P50 CA172012 to L.B), American Society of Hematology (K.L.B), EvansMDS Foundation (K.L.B.), European Hematology Association (E.P.), Gabrielle’s Angels Foundation (E.P.), V Foundation (E.P.), Geoffrey Beene Foundation (E.P), Starr Cancer Consortium (to R.L., A.Z., M.B, R.P.), and the Cancer Colorectal Cancer Dream Team Translational Research Grant (SU2C-AACR-DT22-17 to L.D.). E.P. is a Josie Robertson Investigator. M.M is supported by funds from the Intramural Research Program of the National Cancer Institute, National Institutes of Health. The KoCH cohort was supported through a grant from the Korea Health Technology R&D Project through the Korea Health Industry Development Institute (KHIDI), funded by the Ministry of Health & Welfare, Republic of Korea (grant number: HI14C1277). We thank the Global Science experimental Data hub Center (GSDC) and Korea Research Environment Open NETwork (KREONET) service for data computing and network provided by the Korea Institute of Science and Technology Information (KISTI).Work performed at Memorial Sloan Kettering Cancer Center was supported in part by the Cancer Center Support Grant (grant no. P30 CA008748), the Marie Josee and Henry R Kravis Center for Molecular Oncology, Cycle for Survival, and MSK Molecular Diagnostics Service.

## Extended Data Figures/Tables

**Extended Data Figure 1.**
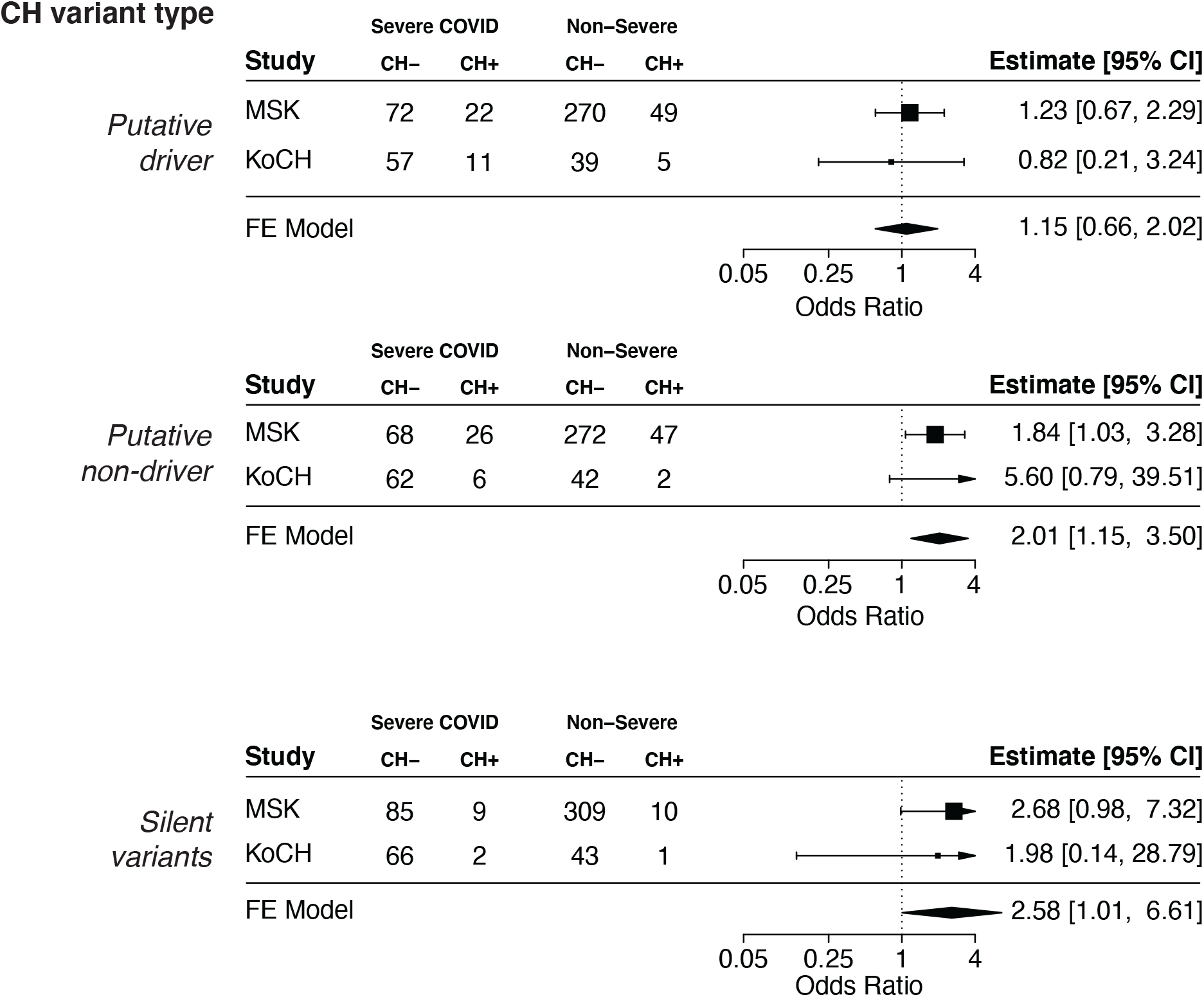
Association between CH and Covid-19 severity. Shown are the results from logistic regression adjusted for age, gender, race, smoking, diabetes, cardiovascular disease, COPD/asthma, cancer primary site (if history of malignancy), exposure to cytotoxic cancer therapy for the MSK and Korea Consortia. Summary statistics for a fixed effects meta-analysis are shown.

**Extended Data Figure 2.**
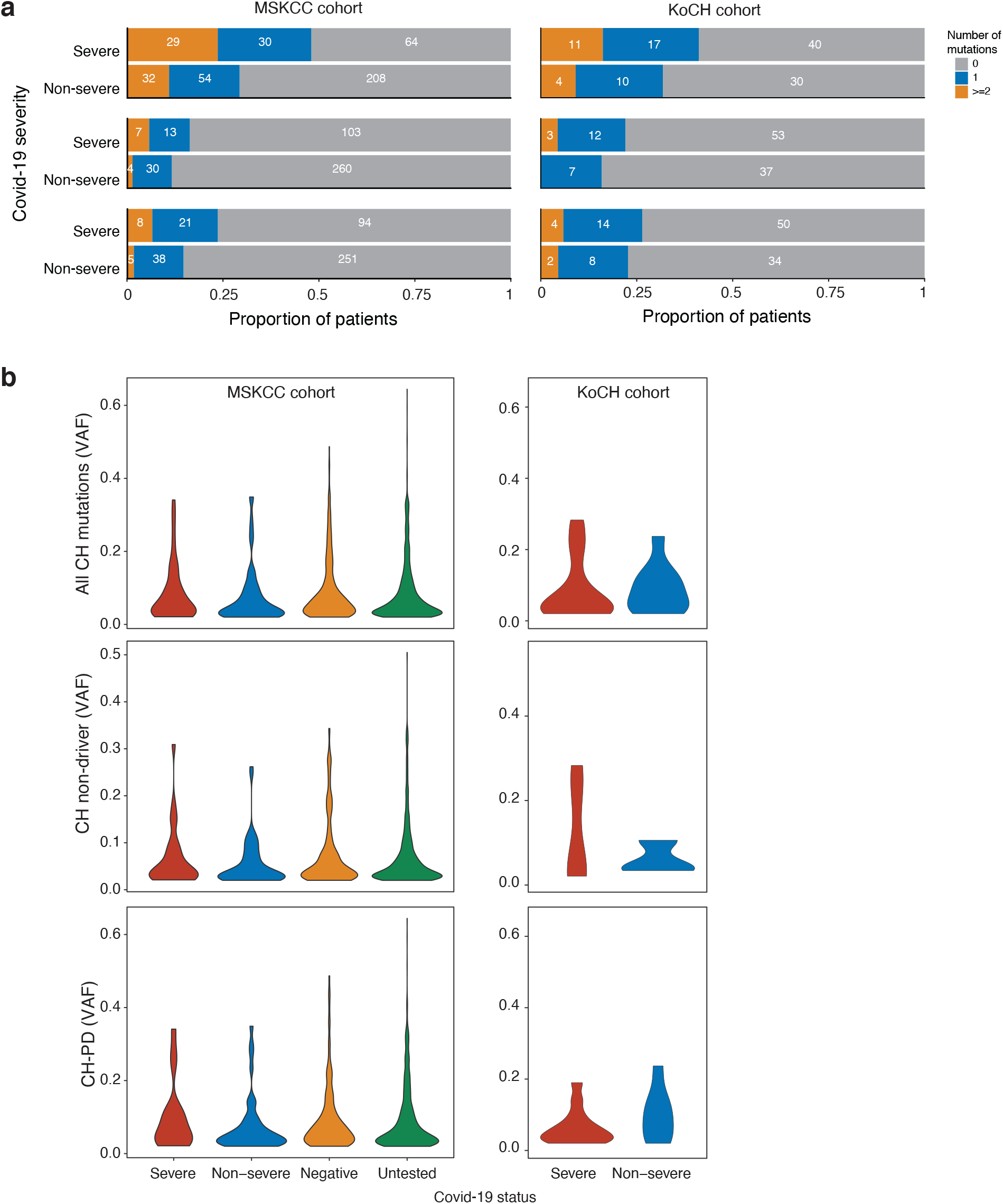
Number of mutations and variant allele fraction of CH by Covid-19 Status. a) Number of CH mutations among those with severe and non-severe Covid-19 b) VAF of CH mutations by Covid-19 severity and infection status.

**Extended Data Figure 3.**
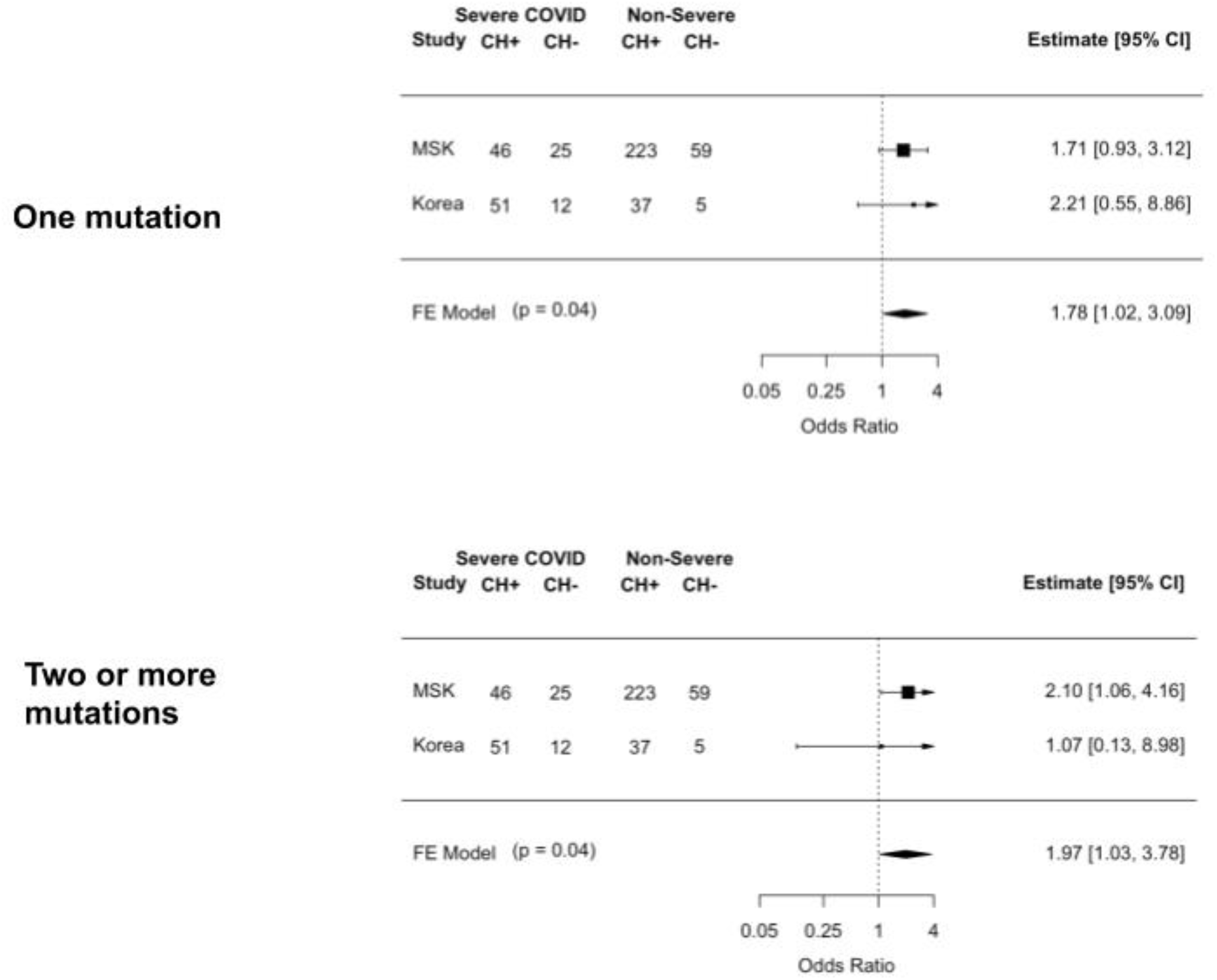
Association between CH and Covid-19 severity stratified by the number of mutations. Shown are the results from logistic regression comparing the odds ratios of severe Covid-19 among those with one mutation and those with two or more mutations. Models were adjusted for age, gender, race, smoking, diabetes, cardiovascular disease, COPD/asthma, cancer primary site (if history of malignancy), exposure to cytotoxic cancer therapy for the MSK and Korea Consortia. Summary statistics for a fixed effects meta-analysis are shown.

**Extended Data Figure 4.**
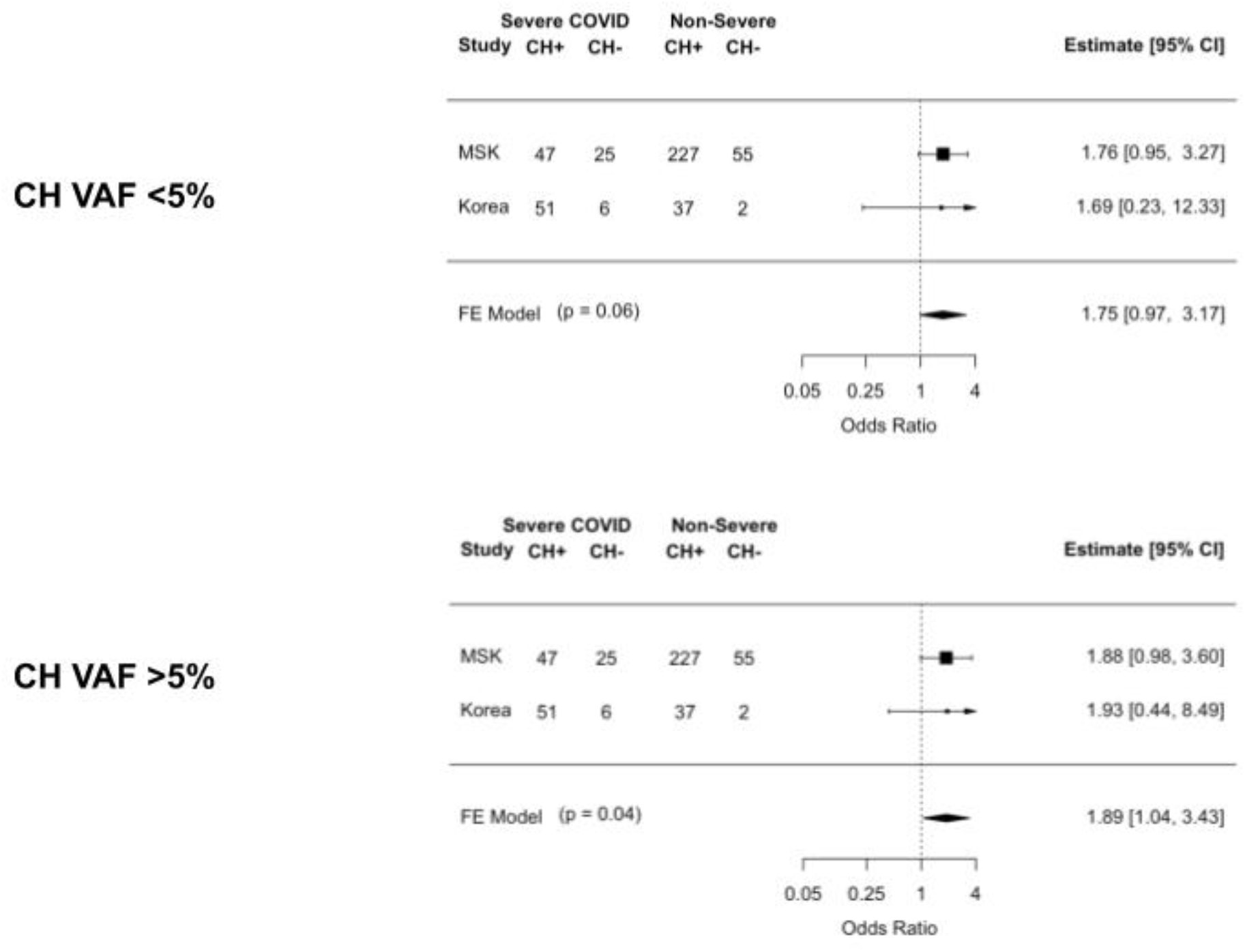
Association between maximum VAF of CH-mutation(s) and Covid-19 severity. Shown are the results from logistic regression comparing the odds ratios of severe Covid-19 among those with one or more CH mutations <5% VAF compared to no CH and CH with a VAF >5% and no CH. Models were adjusted for age, gender, race, smoking, diabetes, cardiovascular disease, COPD/asthma, cancer primary site (if history of malignancy), exposure to cytotoxic cancer therapy for the MSK and Korea Consortia. Summary statistics for a fixed effects meta-analysis are shown.

**Extended Data Table 1.**
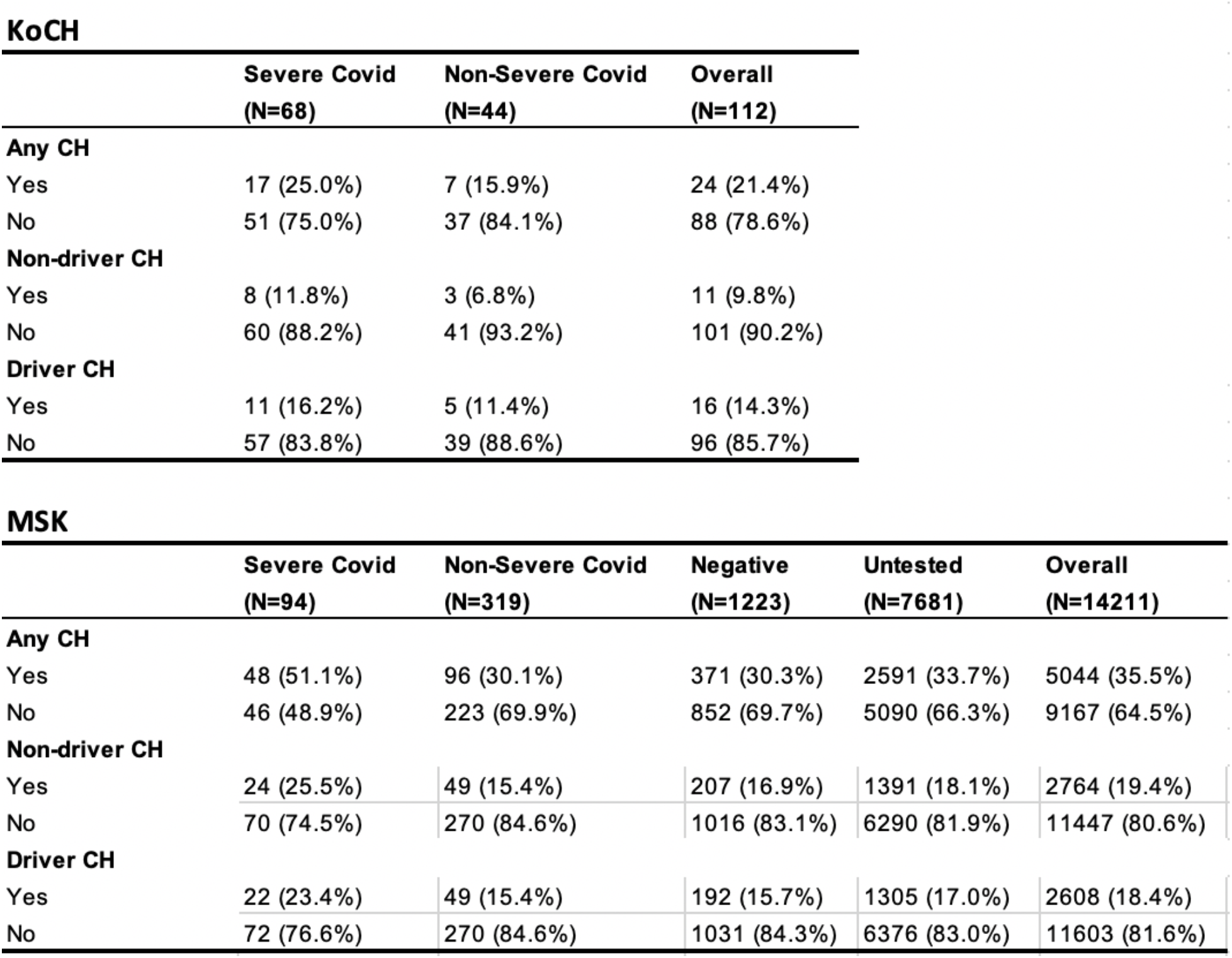
Frequency of clonal hematopoiesis by Covid-19 status.

**KoCH**
|  | **Severe Covid**<br>**(N=68)** | **Non-Severe Covid**<br>**(N=44)** | **Overall**<br>**(N=112)** |
| --- | --- | --- | --- |
| **Any CH** |  |  |  |
| Yes | 17 (25.0%) | 7 (15.9%) | 24 (21.4%) |
| No | 51 (75.0%) | 37 (84.1%) | 88 (78.6%) |
| **Non-driver CH** |  |  |  |
| Yes | 8 (11.8%) | 3 (6.8%) | 11 (9.8%) |
| No | 60 (88.2%) | 41 (93.2%) | 101 (90.2%) |
| **Driver CH** |  |  |  |
| Yes | 11 (16.2%) | 5 (11.4%) | 16 (14.3%) |
| No | 57 (83.8%) | 39 (88.6%) | 96 (85.7%) |

|  | **Severe Covid**<br>**(N=94)** | **Non-Severe Covid**<br>**(N=319)** | **Negative**<br>**(N=1223)** | **Untested**<br>**(N=7681)** | **Overall**<br>**(N=14211)** |
| --- | --- | --- | --- | --- | --- |
| **Any CH** |  |  |  |  |  |
| Yes | 48 (51.1%) | 96 (30.1%) | 371 (30.3%) | 2591 (33.7%) | 5044 (35.5%) |
| No | 46 (48.9%) | 223 (69.9%) | 852 (69.7%) | 5090 (66.3%) | 9167 (64.5%) |
| **Non-driver CH** |  |  |  |  |  |
| Yes | 24 (25.5%) | 49 (15.4%) | 207 (16.9%) | 1391 (18.1%) | 2764 (19.4%) |
| No | 70 (74.5%) | 270 (84.6%) | 1016 (83.1%) | 6290 (81.9%) | 11447 (80.6%) |
| **Driver CH** |  |  |  |  |  |
| Yes | 22 (23.4%) | 49 (15.4%) | 192 (15.7%) | 1305 (17.0%) | 2608 (18.4%) |
| No | 72 (76.6%) | 270 (84.6%) | 1031 (84.3%) | 6376 (83.0%) | 11603 (81.6%) |

## Supplementary Figure/Tables

**Supplementary Figure 1.**
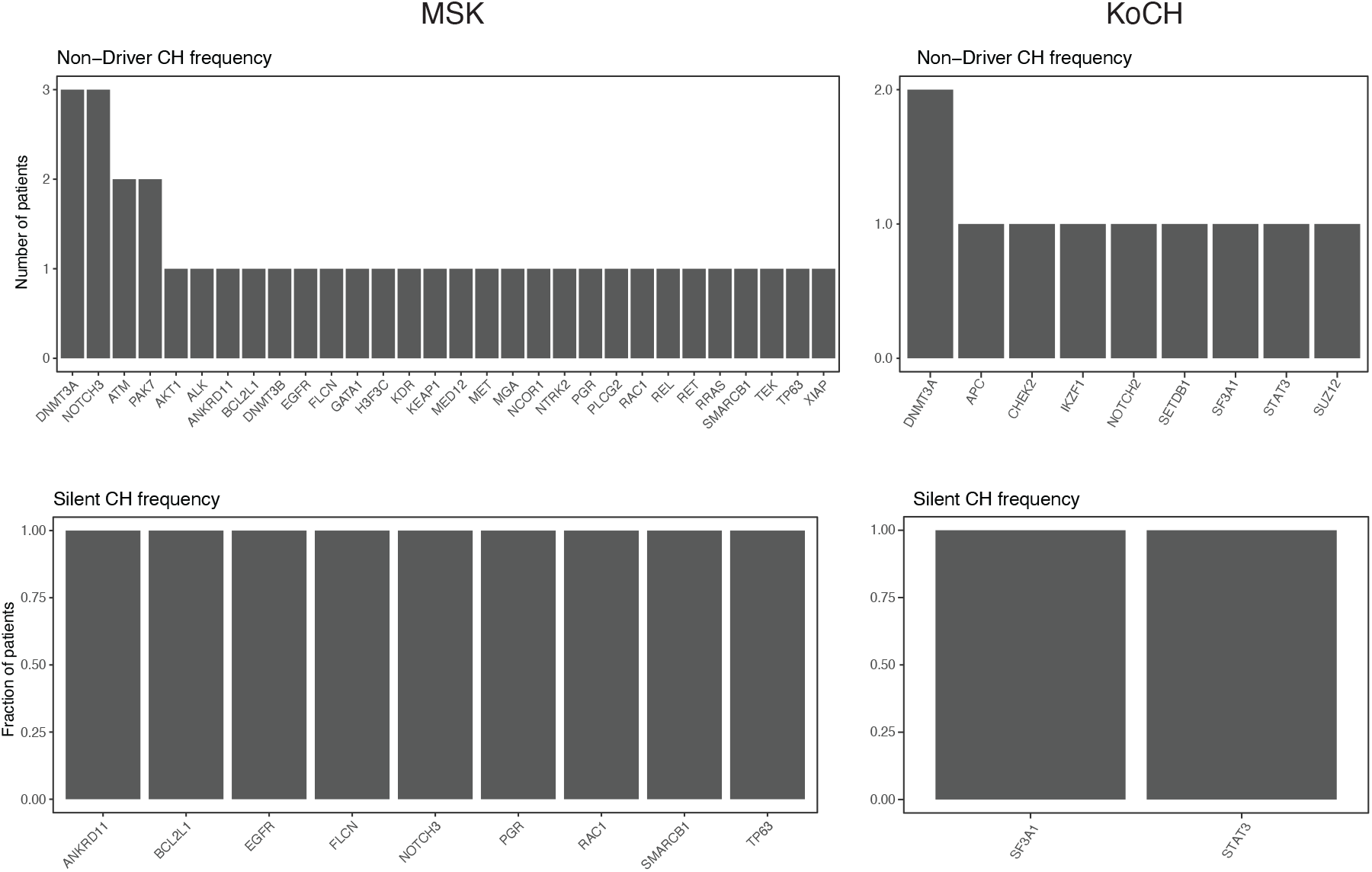
Frequency of genes with non-driver mutations among individuals with severe Covid-19.

## REFERENCES

1. Genovese, G. et al. Clonal Hematopoiesis and Blood-Cancer Risk Inferred from Blood DNA Sequence. N. Engl. J. Med. 371, 2477–2487 (2014).

2. Jaiswal, S. et al. Age-Related Clonal Hematopoiesis Associated with Adverse Outcomes. N. Engl. J. Med. 371, 2488–2498 (2014).

3. Abelson, S. et al. Prediction of acute myeloid leukaemia risk in healthy individuals. Nature 559, 400–404 (2018).

4. Desai, P. et al. Somatic mutations precede acute myeloid leukemia years before diagnosis. Nat. Med. 24, 1015–1023 (2018).

5. Bick Alexander G. et al. Genetic Interleukin 6 Signaling Deficiency Attenuates Cardiovascular Risk in Clonal Hematopoiesis. Circulation 141, 124–131 (2020).

6. Bick, A. G. et al. Inherited causes of clonal haematopoiesis in 97,691 whole genomes. Nature 1–7 (2020) doi:10.1038/s41586-020-2819-2.

7. Del Valle, D. M. et al. An inflammatory cytokine signature predicts COVID-19 severity and survival. Nat. Med. 26, 1636–1643 (2020).

8. Zhou, F. et al. Clinical course and risk factors for mortality of adult inpatients with COVID-19 in Wuhan, China: a retrospective cohort study. The Lancet 395, 1054–1062 (2020).

9. Sano, S. et al. Tet2-Mediated Clonal Hematopoiesis Accelerates Heart Failure Through a Mechanism Involving the IL-1β/NLRP3 Inflammasome. J. Am. Coll. Cardiol. 71, 875–886 (2018).

10. Cai, Z. et al. Inhibition of Inflammatory Signaling in Tet2 Mutant Preleukemic Cells Mitigates Stress-Induced Abnormalities and Clonal Hematopoiesis. Cell Stem Cell 23, 833-849.e5 (2018).

11. Zink, F. et al. Clonal hematopoiesis, with and without candidate driver mutations, is common in the elderly. Blood 130, 742–752 (2017).

12. Poon, G., Watson, C. J., Fisher, D. S. & Blundell, J. R. Synonymous mutations reveal genome-wide driver mutation rates in healthy tissues. bioRxiv 2020.10.08.331405 (2020) doi:10.1101/2020.10.08.331405.

13. Loh, P.-R. et al. Insights into clonal haematopoiesis from 8,342 mosaic chromosomal alterations. Nature 559, 350–355 (2018).

14. Terao, C. et al. Chromosomal alterations among age-related haematopoietic clones in Japan. Nature 584, 130–135 (2020).

15. Coombs, C. C. et al. Identification of Clonal Hematopoiesis Mutations in Solid Tumor Patients Undergoing Unpaired Next-Generation Sequencing Assays. Clin. Cancer Res. Off. J. Am. Assoc. Cancer Res. 24, 5918–5924 (2018).

16. Jaiswal, S. et al. Clonal hematopoiesis and risk for atherosclerotic cardiovascular disease. N. Engl. J. Med. 377, 111–121 (2017).

17. Meisel, M. et al. Microbial signals drive pre-leukaemic myeloproliferation in a Tet2-deficient host. Nature 557, 580–584 (2018).

18. Zeng, H. et al. Antibiotic treatment ameliorates Ten-eleven translocation 2 (TET2) loss-of-function associated hematological malignancies. Cancer Lett. 467, 1–8 (2019).

19. Jee, J. et al. Chemotherapy and COVID-19 Outcomes in Patients With Cancer. J. Clin. Oncol. 38, 3538–3546 (2020).

20. Guan, W. et al. Clinical Characteristics of Coronavirus Disease 2019 in China. N. Engl. J. Med. 382, 1708–1720 (2020).

21. Livingston, E. & Bucher, K. Coronavirus Disease 2019 (COVID-19) in Italy. JAMA 323, 1335 (2020).

22. Petrilli, C. M. et al. Factors associated with hospital admission and critical illness among 5279 people with coronavirus disease 2019 in New York City: prospective cohort study. BMJ 369, (2020).

23. Sung, H. K. et al. Clinical Course and Outcomes of 3,060 Patients with Coronavirus Disease 2019 in Korea, January-May 2020. J. Korean Med. Sci. 35, e280 (2020).

24. Bolton, K. L. et al. Cancer therapy shapes the fitness landscape of clonal hematopoiesis. Nat. Genet. 52, 1219–1226 (2020).

25. Wu, P. et al. Mapping ICD-10 and ICD-10-CM Codes to Phecodes: Workflow Development and Initial Evaluation. JMIR Med. Inform. 7, (2019).

26. Phi, J. H. et al. NPM1 as a potential therapeutic target for atypical teratoid/rhabdoid tumors. BMC Cancer 19, 848 (2019).

